# Prelacteal Feeding and Associated Factors among Mothers of Infants Aged 6 to 12 Months in Chitwan District, Nepal: A Community Based Cross-sectional Study

**DOI:** 10.1101/2021.08.20.21262397

**Authors:** Mani Neupane, Govinda Prasad Dhungana, Bimala Sharma, Dhirendra Paudel

## Abstract

**Background:** A Prelacteal feed is any food except mothers’ milk provided to a newborn before breastfeeding is established. Prelacteal feeding is a major barrier to exclusive breastfeeding and is associated with newborn infection. It is a prevalent practice in Nepal. This study aimed to investigate prelacteal feeding practice and associated factors among mothers of infants aged 6 to 12 months in Chitwan district, Nepal.

**Methods:** A community-based cross-sectional study was conducted. 297 mothers of infants aged 6 to 12 months were selected using a systematic random sampling technique. Descriptive statistics, univariate and binomial logistic regression analysis were done to identify the factors associated with prelacteal feeding practice. Variable with a p-value <0.05 were identified as statistically significant factors.

**Result:** The prevalence of prelacteal feeding was 40.1% (95% CI: 34.5%-45.7%). Formula milk (n=109), animal milk (n=13), Plain water (n=6) were some of the types of prelacteal feed reported. Respondents in the 25-29 years age group were about 58.2% less likely to practice prelacteal feeding (adjusted odds ratio (AOR)=0.415, 95% confidence interval (CI):0.209-0.837) as compared to the respondents belonging to 35 years or more age group. Economic status was another factor associated with prelacteal feeding practice. Those mothers with poorer economic status were about 57.9 % (AOR= 0.421, CI: 0.179-0.992) less likely to practice prelacteal feeding than the mothers belonging to the richest. Similarly, mothers having poor knowledge on prelacteal feeding were found about three (AOR= 2.661, CI: 1.514-4.674) times more likely to have prelacteal feeding than those mothers who had good knowledge about prelacteal feeding. Home delivery and caesarean section in case of institutional delivery were two major reasons stated for providing prelacteal feeding.

**Conclusion:** Prelacteal feeding was commonly practiced in the Chitwan district, Nepal. Mother’s age, economic status, mothers’ knowledge of prelacteal feeding practices, and mode of delivery were factors associated with prelacteal feeding practices. Therefore, awareness and knowledge on the risk associated with prelacteal feeding, promotion of institutional delivery, timely initiation of breastfeeding, and avoidance of prelacteal feeding are important measures for preventing prelacteal feeding practices in Chitwan district, Nepal.

## INTRODUCTION

Mother’s milk is a unique blessing of God that has nourished the complete needs of a newborn which is regarded as the healthiest, the freshest, the safest, the most accessible, and perfect nourishment for a child during the first 2 years of life [1]. Breastfeeding is the normal and ideal way of providing young infants with the nutrients they required for healthy growth and development [2]. Breast milk is a perfect nutrient, easily digested, efficiently used, and protects against infection. Meanwhile, breastfeeding fosters mother and child bonding. Breastmilk aids in the overall development of a child helps delay a new pregnancy and protects the mother’s health. Likewise, also minimize the out-of-pocket expenditure for artificial feeding [3].

Breastfeeding is a well-established and globally suggested intervention for the betterment of child nutrition. Breastfeeding reduces infant and young children deaths and is globally engrave as being the best for any neonate [4]. The World Health Organization (WHO) recommends optimal breastfeeding; that newborns should have early initiation of breastfeeding within one hour after birth (timely initiation of breastfeeding), exclusive breastfeeding, and continued breastfeeding for two years or beyond with the addition of timely, adequate, safe and properly fed complementary foods [5].

Any food like ghee (refined butter), sugar, sugar juice, honey, raw cow/buffalo/goat milk, formula milk, etc. provided to a newborn before the initiation of mother’s breastfeeding is considered a pre-lacteal feed [6]. The practice of pre-lacteal feeding still exists in South Asia Region, which has lesser nutrient and immunological function. Pre-lacteal feeding is one of the main visible barriers to exclusive breastfeeding. The knowledge and information regarding various determinants of pre-lacteal feeding are found most crucial for promoting exclusive breastfeeding and early initiation of breastfeeding [6]. Pre-lacteal feeding impedes suckling and making breastfeeding difficult which establishes a great dispute to ensure optimal breastfeeding and infant nutrition. Most of the infants who receive pre-lacteal feeding were more likely to be stunted and wasted [7]. The risk of illness from acute respiratory tract infection and diarrhea is increased due to pre-lacteal feeding and it is linked with poor breastfeeding outcomes [8]. Prelacteal feeding practices interfere with colostrum feeding as a result it diminishes the immunological benefits a newborn receives from colostrum milk, thus increasing his/her susceptibility to infection. In addition, pre-lacteal feeding can be a direct cause of illness by exposing infants to contaminated feeds, utensils, water, or even a hand [9].

Pre-lacteal feeding is practiced globally in many countries and accounts for 54.6% in Southeast Asia [8].In Nepal, 28% of infants are given a pre-lacteal feed. Pre-lacteal feeding is twice as high in the terai region (37%) than in the mountain region (17%) and hilly region (18%). Among the sub-regions, the highest proportion of children receiving a pre-lacteal feed is observed in the central terai sub-region (53%) [10]. Previous studies conducted in different countries report that several sociodemographic, maternal, and health service and knowledge-related factors significantly correlated with this practice [6,8,11].

Although pre-lacteal feeding is widely practiced in Nepal, to date, only a few studies from Nepal have reported on the factors of pre-lacteal feeding. This study aimed to identify pre-lacteal feeding practice and associated factors among mothers of an infant aged 6-12 months in Chitwan district, Nepal.

## METHODS

### Study setting and participants

This study was conducted in Bharatpur metropolitan city, in the central-southern part of Nepal located in Chitwan district in 2018. A community-based cross-sectional study was conducted among the mothers having 6 to 12 month’s infants. The sample size was determined using a formula for estimation of single population proportion as follows [12]:

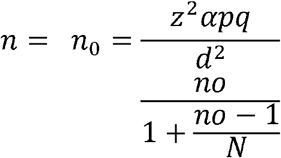

Where, n= the required sample size, Z= critical value for normal distribution at 95% confident level (1.96), p= prevalence of exclusive breastfeeding practice (66%) [13],d= 0.05 (5% margin of error) allowed in the study, and an estimated non-response rate 10%.

#### Sampling procedure

A total of eight wards out of 29 wards of the Metropolitan City was chosen randomly; they were ward number 8, 14, 16, 17, 18, 22, 26, 27. The list of all mothers having 6 to 12 months infant was obtained from the immunization record file of all the Health posts and PHCC of the selected wards. Systematic random sampling was done to select respondents from the obtained sampling frame. The sampling frame for the study was 1270, while the sample size was 299. Sampling frame (1270)/ Sample size (299) = 4.24^th^ interval ∼4^th^ interval. The first sample was chosen randomly from 1 to 4 and the third number was selected, then every 4^th^ name of a mother from the sampling frame obtained from the immunization register was selected as a respondent for the study.

### Measurement of variables

#### Dependent variable

In this study, the outcome variable was prelacteal feeding practice among the mothers having 6 to 12 months infants. The feeds included Formula milk plain water, animal milk, glucose water, honey, ghee, saltwater, and/or fruit juice, etc. It was measured as ‘yes’ if the infants were given foods and or drinks other than mother’s milk before the initiation of breastfeeding and otherwise “no”. The same measurement was used in several previous studies [6-7].

#### Independent variable

The independent variables were socio-demographic and economic characteristics (age of mother, religion, family type, maternal education, occupation, wealth quintile); MCH service utilization characteristics (antenatal care, breastfeeding counseling/information, place of delivery, mode of delivery, and postnatal care), knowledge related characteristics (prelacteal feeding) maternal and infant characteristics (condition of breast, maternal participation in decision making). Wealth quintile was a composite measure of socioeconomic status used in the Nepal Demographic Health Survey (NDHS), 2011[10]. It involves principle component analysis of details of household assets and expenditure. Based on this, a household was categorized into five groups from the poor to the least poor groups corresponding from the lowest to the highest quintiles. Breast related problem was measured by asking the respondents if they had the presence of any one of the following conditions: breast engorgement, crack nipple, breast abscess, retracted nipple, and painful breastfeeding. It was categorized as “yes” and “no”. Maternal illness was measured in terms of any medical conditions of mothers after delivery up to 6 months period.

### Data collection instrument, methods, and process

Data were collected using a pre-tested semi-structured questionnaire adapted from the Nepal Demographic Health Survey 2011, and other similar studies. The adapted questionnaire was modified and contextualized to fit the local situation and the research objective. The questionnaire was prepared first in the English language, was translated to Nepali, and then back into the English language so that the notion of the study remained unchanged. The final Nepali version of the questionnaire was used to collect the data and had ensured that respondents had understood the questions and consistency was maintained during the interview. Face to face interview was conducted with all the mothers selected for study at their homes. All interviews were performed by the principal investigator himself. Consent was obtained from all respondents before conducting the interview.

### Statistical analysis

The Filled questionnaires were checked for correctness & completeness at the end of each interview and thorough crosschecking and editing were done at the end of each day of data collection. Collected data was cleaned, coded, and entered in Epi-Data version 3.1 and was imported to Statistical Package for Social Science (SPSS) IBM SPSS software version 20 (IBM Corp., Armonk, NY, USA) for analysis. Descriptive statistics were computed. Bivariate and multivariate logistic regression analysis was applied to find out the factors affecting the dependent variable. Both Crude and Adjusted odds ratios were presented with 95% confidence intervals. Variable with p-value <0.2 in the binary logistic regression analysis were included in the multivariable logistic analysis. The Hosmer-Lemeshow goodness-of-fit was used to check the model fitness. The model was fit for the variables included as indicated by the p-value.

### Ethical consideration

The study was approved by the Institutional Review Committee (IRC) of Chitwan Medical College (CMC). An official letter was provided from CMC to District Public Health Office. Informed consent was taken from the participants before the interview. The participants were also assured about the confidentiality of the information they provided. Participants’ dignity was maintained by giving them a right to reject or discontinue the study at any time of their choice.

## RESULT

A total of 297 mother-child pairs were included in the study, resulting in a response rate of 99.3%. The mean (± SD) age of respondents was 25.96 years (±4.00) and ranged from 15 to 35 years. A majority (42.1%) of respondents were in the age group of 25 to 29 years. Of the total, 86.5%of the respondents belonged to the Hindu religion; and 61.3% were from joint families. Regarding education level; 57.3% were from secondary and above, and 62.6% were involved in household works followed by agriculture 23.2%. Regarding economic status, one-fifth (20.2%) were from the poorest (Table 1).

**Table 1.** Socio demographic and Economic characteristics (n=297)

| Variables | Number | Percentage |
| --- | --- | --- |
| Age of respondent |  |  |
| 15-19 | 9 | 3.0 |
| 20-24 | 104 | 35.0 |
| 25-29 | 125 | 42.1 |
| 30-34 | 47 | 15.8 |
| ≥35 | 12 | 4.0 |
| Mean age ± SD (In year) | 25.96±4.00 |  |
| Religion of respondent |  |  |
| Hindu | 257 | 86.5 |
| Buddhist | 34 | 11.4 |
| Christian | 5 | 1.7 |
| Muslim | 1 | .3 |
| Type of family |  |  |
| Joint | 182 | 61.3 |
| Nuclear | 115 | 38.7 |
| Education level of mother |  |  |
| Illiterate | 11 | 3.7 |
| Literate | 12 | 4.0 |
| Basic level<sup>a</sup> | 104 | 35.0 |
| Secondary and above level<sup>b</sup> | 170 | 57.3 |
| Occupation of respondent |  |  |
| House maker | 186 | 62.6 |
| Agriculture | 69 | 23.2 |
| Business | 28 | 9.4 |
| Service | 10 | 3.4 |
| Labor | 4 | 1.3 |
| Economic status |  |  |
| Poorest | 60 | 20.2 |
| Poorer | 59 | 19.9 |
| Middle | 59 | 19.9 |
| Richer | 60 | 20.2 |
| Richest | 59 | 19.9 |

<sup>a</sup> includes primary level and basic level, <sup>b</sup> includes secondary, higher secondary and above higher education level

Of the total, 73.9% of respondents had attended 4 or more times ANC visits; 97.6% of respondents delivered their baby in a health institution, of them, 77.1%were normal delivery, and 22.9% were cesarean sections. Regarding PNC visits, 97.6% of the respondent had done at least one PNC visit. Among the total, 53.9% of mothers had received information on breastfeeding, and of them, 51.3% of respondents were counselled on avoidance of prelacteal feeding. Prevalence of prelacteal feeding was 40.1% (95% CI: 34.5%-45.7%). Among respondents, 53.8% reported caesarean section as the main cause behind prelacteal feeding practice. Prelacteal feedings provided to the baby were formula milk 91.6%, animal milk 10.92%, and plain water 5.0% respectively. Regarding knowledge on Prelacteal feeding (44.4%) had poor knowledge followed by fair knowledge 30.6%, no knowledge 17.5%, and good knowledge 7.4% (Table 2).

**Table 2.** MCH services utilization and prelacteal feeding.

| Variables | Number | Percentage |
| --- | --- | --- |
| Number of ANC visit (n=295) |  |  |
| < 4 | 77 | 26.1 |
| ≥ 4 | 218 | 73.9 |
| Place of delivery |  |  |
| Health institution | 290 | 97.6 |
| Home | 7 | 2.4 |
| Mode of delivery |  |  |
| Normal vaginal delivery | 229 | 77.1 |
| Cesarean section | 68 | 22.9 |
| Delivery attendant |  |  |
| Health professionals | 290 | 97.6 |
| Relatives/friends | 6 | 2.0 |
| FCHVs | 1 | 0.3 |
| Number of PNC visit (n=297) |  |  |
| 0 | 7 | 2.4 |
| 1 | 216 | 72.7 |
| 2 | 27 | 9.1 |
| 3 | 47 | 15.8 |
| Counseling /information on breastfeeding (n=297) |  |  |
| Yes | 160 | 53.9 |
| No | 137 | 46.1 |
| Counseling received areas (n=160) |  |  |
| Practice exclusive breastfeeding | 123 | 76.9 |
| Colostrum should be feed | 102 | 63.8 |
| Prelacteal feeding should be avoided | 82 | 51.3 |
| Early initiation of breastfeeding | 57 | 35.6 |
| Others | 6 | 3.8 |
| Prelacteal feeding (n=297) |  |  |
| No | 178 | 59.9 |
| Yes | 119 | 40.1 |
| Types of Prelacteal feeding (n=119) |  |  |
| Formula milk | 109 | 91.6 |
| Animal milk | 13 | 10.92 |
| Plain water | 6 | 5.0 |
| Reason for Prelacteal feeding (n=119)* |  |  |
| Caesarean section | 64 | 53.8 |
| Delay milk secretion | 53 | 44.5 |
| Health worker advised | 43 | 36.1 |
| Maternal illness | 42 | 35.3 |
| Baby not able to suck mother's milk | 8 | 6.7 |
| Lack of information | 4 | 3.4 |
| Others | 5 | 4.2 |
| level of knowledge on prelacteal feeding |  |  |
| No knowledge | 52 | 17.5 |
| Poor knowledge | 132 | 44.4 |
| Fair knowledge | 91 | 30.6 |
| Good knowledge | 22 | 7.4 |

\* multiple response

In the study, 11.4% of respondents had suffered from illness during the postpartum period of 0 to 6 months and 29.0% of respondents had faced breast-related problems. Of them, 75.3% had faced painful breastfeeding (Table 3).

**Table 3.** Maternal and Infant related factors, Chitwan, Nepal, 2018.

| Variable | Number | Percent |
| --- | --- | --- |
| Breast related problem(n=297) |  |  |
| No | 211 | 71.0 |
| Yes | 86 | 29.0 |
| Types of Breast related problem(n=86) |  |  |
| Painful breastfeeding | 64 | 75.3 |
| Crack nipple | 34 | 40.0 |
| Brest engorgement | 31 | 36.5 |
| retracted nipple | 8 | 9.4 |
| Others | 4 | 4.7 |
| Level of participation in decision making (n=297) |  |  |
| No participation | 127 | 42.8 |
| Moderate participation | 38 | 12.8 |
| High participation | 132 | 44.4 |
| Newborn weight at birth (n=297) |  |  |
| <2500 | 18 | 6.1 |
| ≥2500 | 279 | 93.9 |

\*multiple response

There was a total of 297 cases for analysis. Due to the presence of outliers, 2 cases were excluded from analysis out of total cases to fit the model adequacy. To avoid multicollinearity, variable (PNC utilization) was excluded from the final analysis. Likewise, another variable (modes of delivery) was excluded from the final analysis because of less number frequency in any one cell in order to fit the model. Respondents found in the 25-29 years age group were about 58.2 percent less likely to practice prelacteal feeding (AOR=0.415, CI:0.209-0.837) than respondents belonging to the 35 or more age group. The result showed that a mother’s age is one of the factors for prelacteal feeding. In the case of economic status, poorer respondents were about 57.9 percent (AOR= 0.421, CI: 0.179-0.992) less likely to practice prelacteal feeding than those respondents who belonged to the richest. This finding showed that economic status also plays a significant role in prelacteal feeding practice. Similarly, the relationship between knowledge and prelacteal feeding was also found significantly associated after controlling all other variables with an adjusted odds ratio of 2.661. This showed that respondents who had poor knowledge of prelacteal feeding were found about three (AOR= 2.661, CI: 1.514-4.674) times more likely to have prelacteal feeding than those respondents who had good knowledge about prelacteal feeding (Table 4).

**Table 4.** Multivariate analysis of factors associated with prelacteal feeding.

| **Characteristics** | **Unadjusted OR**<br>**(95% CI)** | **Adjusted OR**<br>**(95% CI)** | ***p***<br>**value** |
| --- | --- | --- | --- |
| Age of Mother |  |  |  |
| 15-19 | 1.933(0.441-8.467) | 2.050(0.402-10.447) | 0.388 |
| 20-24 | 0.534(0.279-1.022) | 0.493(0.239-1.015) | 0.055 |
| 25-29 | 0.472(0.251-0.888) | 0.418(0.209-0.837) | 0.014 |
| 35 or more | 1 | 1 |  |
| Religion |  |  |  |
| Hindu | 1 | 1 |  |
| Non Hindu <sup>b</sup> | 2.192(1.118-4.297) | 1.999(0.934-4.240) | 0.071 |
| Main occupation |  |  |  |
| House maker | 1 | 1 |  |
| Business | 1.692(0.758-3.775) | 1.944(0.804-4.703) | 0.14 |
| Agriculture | 1.790(1.021-3.138) | 1.730(0.911-3.827) | 0.094 |
| Paid worker<sup>d</sup> | 1.085(0.349-3.373) | 2.393(0.673-8.510) | 0.178 |
| Economic status |  |  |  |
| Poorest | 0.907(0.438-1.874) | 0.690(0.296-1.606) | 0.389 |
| Poorer | 0.603(0.258-1.276) | 0.421(0.179-0.992) | 0.048 |
| Middle | 0.811(0.389-1.688) | 0.565(0.244-1.312) | 0.184 |
| Richer | 0.683(0.327-1.430) | 0.532(0.231-1.227) | 0.139 |
| Richest | 1 | 1 |  |
| ANC visit |  |  |  |
| <4 | 1.466(0.866-2.483) | 1.117(0.598-2.083) | 0.729 |
| ≥4 | 1 | 1 |  |
| Place of delivery |  |  |  |
| Home | 3.860(0.736-20.232) | 3.958(0.655-23.904) | 0.134 |
| Health institution | 1 | 1 |  |
| Level of knowledge on prelacteal feeding |  |  |  |
| Poor knowledge | 2.592(1.560-4.307) | 2.661(1.515-4.675) | 0.001 |
| Good knowledge | 1 | 1 |  |
| Mother participation in decision making |  |  |  |
| Low participation | 1.875(1.164-3.022) | 1.696(0.963-2.968) | 0.067 |
| High participation | 1 | 1 |  |

1-Reference category, OR- odds ratio, CI- confidence interval. <sup>b</sup> include Buddhist , Christian, Muslim. <sup>d</sup> includes labor, service.

## DISCUSSION

The study revealed Prelacteal feeding practices and their correlates. The study found a high prevalence of prelacteal feeding in the study area; and the predictors were the age of the mother, economic status, and level of knowledge.

In this study, prelacteal feeding included formula milk, animal milk, and plain water was given to 40% of children. The result is consistent with a cross-sectional study conducted among 352 recently delivered women of rural Uttar Pradesh, India by Manas Pratim et al. (40.1%) [11] Another community-based cross-sectional study was conducted selecting 630 mothers of children aged less than 24 months by systematic random sampling technique in Raya Kobo district, North Eastern Ethiopia by Misgan et al. showed 39% prevalence of Prelacteal feeding [7]. However, Nepal demographic and health survey 2011 finding shows prelacteal feeding of 28% which is lower than this study. Similarly, another study was done in Nepal by Khanal et al. also showed Prelacteal feeding was 26.5% which was again lower than this study [10,6]. Furthermore, a study conducted by Shrivastava et al among 508 mothers of infants attending health centers in eastern Nepala reported that Prelacteal feeding was 17.9 which was much lower than our study finding [14]. The practice of prelacteal feeding seems to be higher in the study area while comparing the finding with previous national and subnational studies. In spite of following the same definition of prelacteal feeding, the discrepancy in the practice might be area-related or other specific reason prevailing in the study area that requires further studies.

In the international context, the prevalence of this practice was 11.1% in a study conducted by Nigus et al. at Northen Ethiopia and 26.8% in the study conducted by Amare et al [8, 15]. Children were found given prelacteal feeds. In contrast to the study, a cross-sectional study from Vietnam showed 73.3%of the newborns were fed prelacteal which was higher than present study findings [9]. Likewise, another study conducted by Gupta et al. in Delhi reported that 47.4% of the newborns were fed prelacteal [16]. The study found mixed types of results regarding the practice of prelacteal feeding.

The study found age inversely associated with the practice of prelacteal. Younger mothers were less likely to feed Prelacteal to their newborns. A study conducted in Northern India by Manas R et al. had also found a significant relationship between the practice of prelacteal feeding and the age of the mother [11].

In the study, the economic status of the respondents was significantly correlated with the prelacteal feeding of the newborns. The respondents from poorer classes were less likely to feed Prelacteal than the respondents belonging to the richest group. The study conducted by Khanal et al. findings from the Nepal Demographic and Health Survey 2011 showed that the mothers from the middle wealth quintile were more likely to provide their children with prelacteal feeds than their counterparts. The lower socio-economic groups may have less access to the expensive prelacteal feeds such as ghee or honey, formula milk and therefore exclusive breastfeeding is the only option available to them [6]. A study conducted by Tariku et al. had shown a contrasting result that respondents being in the lowest wealth status increases the odds of prelacteal feeding by 2 times [8].

In the study, the mother’s knowledge about prelacteal feeding was one of the significant factors associated with prelacteal feeding. This finding is consistent with the cross-sectional study done in Raya Kobo district, North Eastern Ethiopia by Legesse et al. [7]. Similarly, a community-based cross-sectional study conducted by Tariku et al. found higher odds of prelacteal feeding among mothers with poor knowledge of infant and young child feeding (IYCF) compared to mothers with the highest knowledge of IYCF [8].

The mode of the delivery was found one of the important determining factors of prelacteal feeding. As there was one hundred percent prelacteal feeding in the CS delivery, the mode of delivery could not be included in the integrated logistic model. A study conducted on Ghanaian women showed that respondent respondents who had done caesarean delivery were around two (AOR= 2.46, CI: 1.55-3.90) times more likely to provide prelacteal feed to their newborn than the respondent having a vaginal delivery. The findings from this study indicate that cesarean delivery was significantly associated with a higher likelihood of prelacteal feeding. Furthermore, Petal *et al*. (2013) and Raheem et al. (2014) both identified a cesarean delivery to be a significant risk factor for prelacteal feeding [17].

The study revealed that some reasons for prelacteal were lack of appropriate breastfeeding interventions in Vietnam, particularly during routine health care healthcare, lack of knowledge and confidence of mothers through appropriate perinatal counseling and support, poor routine ANC, and post-delivery management. Conclusively, overall variation in the prevalence of prelacteal feeding within Nepal, within south Asian countries, and other developing countries were likely due to variation in socio-demographic and economic status, MCH services utilization, modes of delivery, place of delivery, counseling, and information on breastfeeding practices and urban setting.

## CONCLUSION

This study was a community-based cross-sectional study aimed at highlighting the association of some key factors to Prelacteal feeding practices and it was found commonly practiced in the Chitwan district. Age of mother, economic status, and mother’s knowledge on prelacteal feeding practices were important factors of Prelacteal feeding practice. Awareness and knowledge on the risk associated with prelacteal feeding, promotion of institutional delivery, and timely initiation of breastfeeding is an essence to reduce its practices as well as a concerned authority should ensure no prelacteal feeding even in case of cesarean section delivery.

## Data Availability

The correspondent authors can provide the data on reasonable requests.

## Authors’ contribution

MN conceived the concept of study design, did data collection, and performed the statistical analysis. MN with significant contribution from BS prepared a draft of the manuscript.GD contributed to the reanalysis of the data and critically revising the manuscript. BS contributed to revising the manuscript with a significant intellectual contribution. All of the authors contributed to revision and agreed on the final version of the manuscript. DP supervised the manuscript preparation.

## Acknowledgement

The authors would like to thank all the respondents who devoted their valuable time to the study and provided the information. The authors also sincerely acknowledged Female Community Health Volunteers and Health Post workers supporting in identifying respondents.

## Declaration

The correspondent authors can provide the data on reasonable requests.

